# Adherence to Public Health Recommendations, Restrictions, and Requirements among Priority Populations at Risk for COVID-19 Mortality and Infection in Australia

**DOI:** 10.64898/2026.02.15.26346356

**Authors:** Shanti Narayanasamy, Aimée Altermatt, Anna L. Wilkinson, Katherine Heath, Katherine B. Gibney, Margaret Hellard, Alisa Pedrana

**Author notes:** Corresponding Author Shanti Narayanasamy. These authors contributed equally to this work (Margaret Hellard, Alisa Pedrana).

## Abstract

**Objective:** To examine adherence to COVID-19 public health measures among culturally and linguistically diverse (CALD) and low socio-economic status (SES) populations in Victoria using a unique longitudinal cohort.

**Study Design:** The Optimise Study was a mixed-methods longitudinal cohort and social networks study (September 2020 – December 2023) assessing the impact of COVID-19 and related public health measures in Victoria, Australia. We used a serial cross-sectional design to analyse adherence to public health recommendations, restrictions, and requirements.

**Settings, participants:** The study examines two 28-day periods during the COVID-19 pandemic in Victoria: April 23– May 20, 2021 (‘non-lockdown’), and September 13–October 10, 2021 (‘lockdown’). We explored adherence to three categories of COVID-19 public health measures — Recommendations (non-enforced, longer-term), Restrictions (mandated during lockdown periods), and Requirements (mandated, longer-term) — among participants who completed questionnaires during these periods. Participants were grouped as: 1) non-CALD high SES (did not meet CALD or low-SES criteria), 2) CALD, or 3) non-CALD low-SES.

**Main outcome measures:** Primary outcomes were adherence to Recommendations, Restrictions, and Requirements during the two study periods.

**Results:** Of 782 participants recruited, 579 (75%) completed a survey or diary during at least one study period and were included in the analysis. Of these, 275 (47%) were in the ‘non-CALD high-SES’ group, 114 (20%) in the CALD group, and 190 (33%) in the ‘non-CALD low-SES’ group. Across all groups, risk-reduction behaviours increased during the lockdown. CALD participants showed higher adherence to some Recommendations and Restrictions compared to the other groups. Overall, 28% left home while awaiting a COVID-19 test result, commonly due to work.

**Conclusions:** High adherence among CALD and ‘non-CALD low-SES’ groups suggest structural barriers, rather than behavioural non-compliance, contributed to higher COVID-19 impacts, highlighting the need for tailored support. During future public health emergencies, better supports are needed for individuals working outside of home to remain in isolation while awaiting a test result.

**Summary box:** **What is already known about this subject?** In Australia, priority populations such as culturally and linguistically diverse (CALD) and low socio-economic status (SES) groups experienced higher COVID-19 infection, mortality and a disproportionate impact from public health restrictions.
**What does this study add?** CALD populations had an overall higher level of adherence to public health behavioural measures during both lockdown and non-lockdown periods compared to non-CALD populations. Over 25% of participants did not comply with stay-at-home requirements while awaiting a COVID-19 test result, largely due to work responsibilities.
**How might this impact on clinical practice?** Pandemic preparedness efforts should focus on understanding the reasons for non-adherence with isolation requirements and considering tailored support during future pandemics to address the diverse

## BACKGROUND

Priority populations during the COVID-19 pandemic in Australia have been defined as those groups who experienced an inequitable burden of disease and disparities in social and economic outcomes during the COVID-19 pandemic.^1,2^ These differences arise due to inequities in the social determinants of health, including education, employment, socio-economic group, housing stability, access to healthcare, and experiences of racism.^1^ Individuals may also experience intersecting layers of inequity and face disproportionate impacts from pandemic response measures.^1^ In Australia, people born overseas, those living in areas of higher socioeconomic disadvantage, and Aboriginal and Torres Strait Islander peoples died from COVID-19 at higher rates than others.^3,4^ Public health restrictions also impacted priority populations disproportionately, specifically culturally and linguistically diverse (CALD) communities, people with disabilities, people experiencing homelessness, children, and individuals in residential aged care.^1^

In pandemics, non-pharmaceutical interventions based on behaviour change are often the first measures to reduce transmission. Understanding adherence during COVID-19 can help predict behaviour in future pandemics. Australian cross-sectional surveys from 2020 indicated high self-reported compliance with public health restrictions, 83% in one survey (N=1,691; national quota sample)^5^ and 79% in another (n=1,595; national sample).^6^ Both found higher adherence among older adults and women, with no associations with other demographic factors such as country of birth or educational attainment.

Few studies have explored adherence to COVID-19 prevention strategies in Australia among priority populations. Findings on the association between socio-economic status (SES) and adherence to public health measures are mixed: one study found a small but positive association between adherence and higher SES,^7^ another found that higher SES predicted lower intentions to comply with public health measures.^8^ Another Australian study (n=323) examining self-reported adherence to three key measures (physical distancing, self-quarantine if unwell, and testing for SARS-CoV-2 with symptoms) found no effect from SES but lower adherence among men and young adults.^9^ These inconsistencies may reflect differences in study populations and outcome definitions.

Evidence on adherence to public health measures among CALD populations in Australia is limited. A 2020 study of older CALD adults in South Australia (n=155) identified high compliance with hand hygiene and avoiding public places, but lower adherence to staying home or avoiding at-risk groups.^10^ Significant gaps remain in understanding behavioural uptake in priority populations.

The study aims to describe adherence to public health recommendations (non-enforced, longer-term), restrictions (mandated during lockdowns) and requirements (mandated, longer-term) among two priority populations at-risk for COVID-19 in Victoria, Australia: low-SES groups and CALD communities. It draws on a unique longitudinal cohort of participants recruited early in the pandemic, and followed beyond the end of emergency measures.

## METHODS

### Study design

The Optimise Study (Optimise) was a mixed-methods longitudinal cohort and social networks study (September 2020–December 2023) that aimed to assess the impact of the COVID-19 pandemic and public health measures on the community in Victoria, Australia; a detailed description of the study is available elsewhere.^11^ We used a serial cross-sectional study design to understand adherence in the Optimise cohort to public health restrictions, recommendations and requirements.

### Setting

During the period that Victoria was under a State of Emergency (March 20, 2020 – December 21, 2021) three broad categories of public health measures were enacted to control the transmission of SARS-CoV-2.^12^ *Recommendations* were public health measures that were not enforced, such as use of hand sanitizer, disinfecting surfaces/objects and social distancing in public (**Table 1**). *Restrictions* were mandated measures including: stay-at-home orders, closure of schools and non-essential businesses, limitations on social gatherings, curfews and travel restrictions. *Requirements* included mandated testing for SARS-CoV-2 for individuals with flu-like symptoms or those who were considered COVID-19 contacts, and isolation of individuals who were symptomatic or awaiting a COVID-19 test result.

**Table 1.**
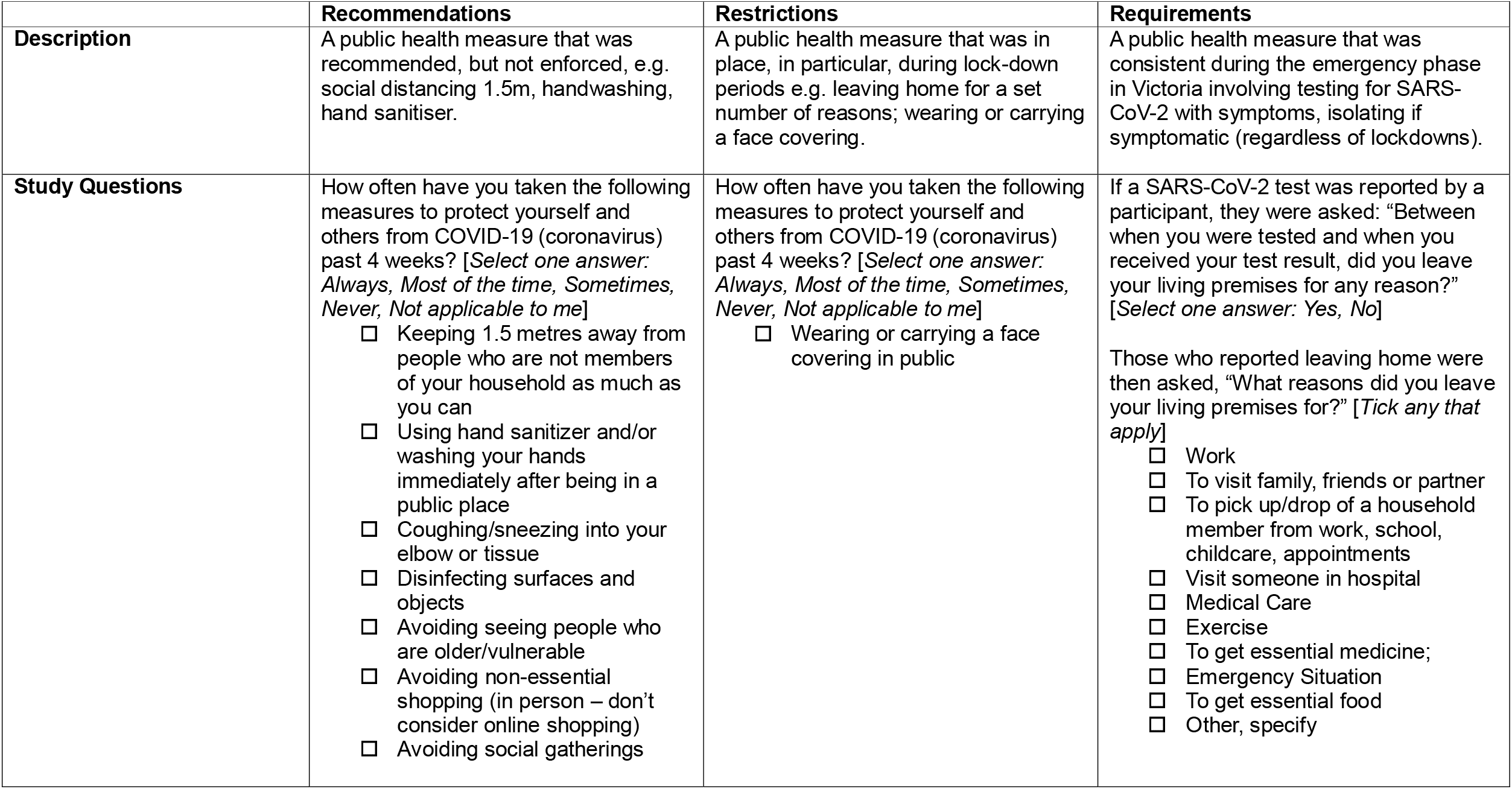
Restrictions, Recommendations and Requirements in Victoria during the COVID-19 State of Emergency.

This study focusses on two distinct 28-day periods: April 23 to May 20, 2021 (‘non-lockdown period’), and September 13 to October 10, 2021 (‘lockdown period’). During the ‘non-lockdown period’ the community transmission of COVID-19 infection in Victoria was low and there were no public health Restrictions in place, however public health Recommendations and Requirements remained (**Figure 1**). During the ‘lockdown period’ the incidence of COVID-19 was high and public health Restrictions, Recommendations and Requirements were in place.

**Figure 1.**
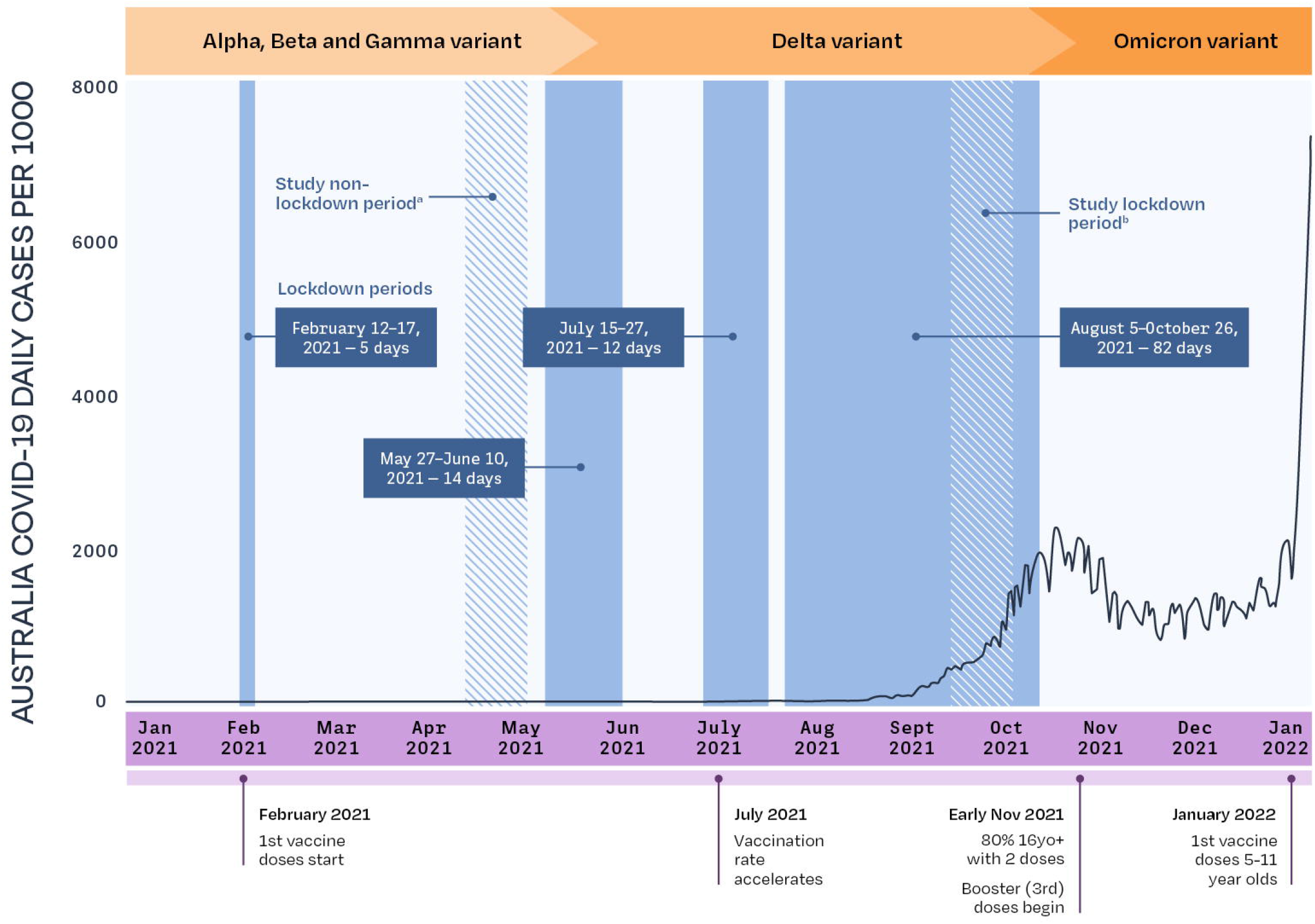
Incidence of SARS-CoV-2 in Victoria, 2021 and public health measures^23,24^. Note: ^a^Non-lockdown period April 23-May 20, 2021 included Recommendations (handwashing and hand hygiene, 1.5m social distancing between individuals from different households) and Requirements (mandatory COVID-19 testing with symptoms and mandatory contact tracing for infective contacts). ^b^Lockdown period September 13-October 10^th^ 2021 included Recommendations and Requirements (as above) and the following Restrictions: curfew from 9pm-5am, mandatory mask use indoors and outdoors in public/shared spaces, remote schooling (exceptions for some children), one visitor per household per day, no travel beyond metropolitan Melbourne, stay-at-home orders (exceptions for essential workers, obtaining essential supplies and 1hr exercise daily), non-essential businesses closed.^25^

This serial cross-sectional study design considered a subset of the data collected throughout the Optimise study period. We included surveys and diaries completed between 23 April and 20 May 2021 (inclusive) for the non-lockdown period, and between 13 September and 10 October 2021 (inclusive) for the lockdown period. These periods were purposively chosen to span 28 days to avoid repeated survey measures from participants, as Optimise participants were requested to complete a survey every 28 days. All participants who contributed a follow-up survey or follow-up diary in either of the two study periods were included in the study.

### Participants

The Optimise study intentionally oversampled people considered at high risk for: a) contracting COVID-19, b) developing severe COVID-19, and c) experiencing negative impacts of government restrictions introduced to reduce SARS-CoV-2 transmission.^11^ Participants were eligible to participate in Optimise if they were aged ≥18 years, resided in Victoria and had access to the internet or a phone. All participants were reimbursed for their participation with monthly electronic gift vouchers.

#### Inclusion criteria

To be included in this study, participants had to have 1) completed the baseline survey, 2) reported a Victorian postcode in their baseline survey, and 3) completed at least one survey or one diary in either of the study time periods.

## Data sources

This study uses three data sources from Optimise: 1) a baseline survey (demographics); 2) follow-up surveys (monthly surveys completed every 28 days collecting similar information as the baseline survey, with past four weeks recall); and 3) follow-up diaries (collected up to four times during one month to capture data on COVID-19 testing, symptoms and mental health in the past seven days).

### Populations (Exposure)

We classified our study population into three groups: 1) ‘non-CALD high-SES’, 2) CALD, and 3) ‘non-CALD low-SES’. Participants were classified in the CALD population if a) their country of birth was not Australia, the United Kingdom or Ireland, or b) their language spoken at home was not English. The CALD definition used for this study reflects the population in Australia with the highest mortality from COVID-19.^13^

Socio-economic groups were defined based on the SEIFA Index of Relative Socio-Economic Disadvantage (IRSD) based on postcode.^146^ The low-SES group was defined as a person residing in an area classified in IRSD deciles 1–6, and high-SES group by IRSD deciles 7– 10 (self-reported data at baseline).

The three groups (‘non-CALD high-SES’, CALD, and ‘non-CALD low-SES’) were mutually exclusive. Allocation was hierarchical: CALD first, then low-SES. Participants meeting criteria for both were assigned to the CALD group only. Those meeting neither were classified as ‘non-CALD high-SES’.

### Behavioural Measures (Outcome)

The primary outcomes were self-reported adherence to public health Recommendations, Restrictions and Requirements during the two study periods of interest (‘lockdown’ and ‘non-lockdown’) (**Table 1**).

Restrictions and Recommendations were assessed using Likert-scale questions on the frequency of risk reduction behaviours (always, mostly, sometimes, never, not applicable) (**Table 1**). Likert scale responses were dichotomised: “always” and “mostly” together, and “sometimes” and “never” together. Responses of “not applicable to me” were classified as missing and were not included in analysis for that question. ‘Not applicable’ responses were treated as missing but did not exclude participants from contributing to other items. Adherence was reported by population group and time period.

The Requirements measure was defined as staying at home between SARS-CoV-2 testing and receiving results. Participants who reported testing for SARS-CoV-2 in the diary recall period (seven days) were asked “Between when you were tested and when you received your test result, did you leave your living premises for any reason?” or for those who had not received their result, “Between when you were tested and now, have you left your living premises for any reason?” Participants who responded “yes” to either question were classified as leaving home between testing and receiving the result. Participants were then asked to select reasons for leaving from a list of options, with multiple responses allowed (**Table 1**).

### Characteristics (Covariates)

Covariates included age, gender, chronic health conditions, location in Victoria (metropolitan or rural), citizenship/permanent residency status, religion, highest level of educational attainment and income. These variables were all measured at baseline. We stratified our analyses by period of survey completion (non-lockdown period, lockdown period).

### Statistical methods

We used descriptive statistics to report on adherence to Recommendations and Restrictions and compare proportions in each group using Chi-squared tests, and reported the associated p-values. We compared differences in proportions using Chi-squared tests between the three exposure groups, for each of the two study periods.

#### Diary data - Requirements

To describe adherence to the isolation requirements, participants were included in the sub-analysis if they reported testing for SARS-CoV-2 in at least one diary in either study period. We report among those who left home while waiting for a result and the reasons given for leaving home. We calculated proportions of participants leaving home after testing (number who reported leaving home divided by number who reported testing) by population group.

#### Sub-group Analysis

We conducted a sub-group analysis to explore the impact of socio-economic status among the CALD group. We divided the CALD group into CALD-high-SES and CALD-low-SES, following our previous definition for high- and low-SES detailed above. The analysis was repeated with four groups: 1) CALD-high-SES, 2) CALD-low-SES, 3) non-CALD low-SES and 4) non-CALD high-SES. We used a Chi-squared test to explore differences.

A p-value of 0.05 was the cut-off for statistical significance.

Data were analysed in R version 4.4.1.

### Patient and Public Involvement

COVID-19 patients and members of the public were involved in this research, a detailed description is noted elsewhere.^11^

### Ethics

Ethics approval for Optimise was provided by the Alfred Human Research Ethics Committee, Approval Number 333/20.

## RESULTS

Of a total 782 participants recruited in the Optimise study, 779 (99%) completed a baseline survey and 773 (99% of 779) reported a Victorian postcode at baseline. A total of 579 (75% of 773) participants completed a survey or a diary in either period and were included in the study.

Of the 579 included participants, 378 (65%) completed a survey during the ‘non-lockdown’ period (29 March – 25 April 2021) and 529 (91%) completed a survey during the ‘lockdown period’ (13 September – 10 October 2021). Three-hundred and forty-nine participants (60%) completed surveys in both study periods.

Four hundred and one participants (69% of 579) completed a diary in the ‘non-lockdown’ period, and 558 (96%) completed a diary in the ‘lockdown’ period. A total of 387 participants (67%) completed at least one diary in both study periods. Of the 579 participants included in the analysis, 275 (47%) were classified into the ‘non-CALD high-SES’ group, 114 (20%) in the CALD group and 190 (33%) in the ‘non-CALD low-SES’ group (**Table 2**).

**Table 2.**
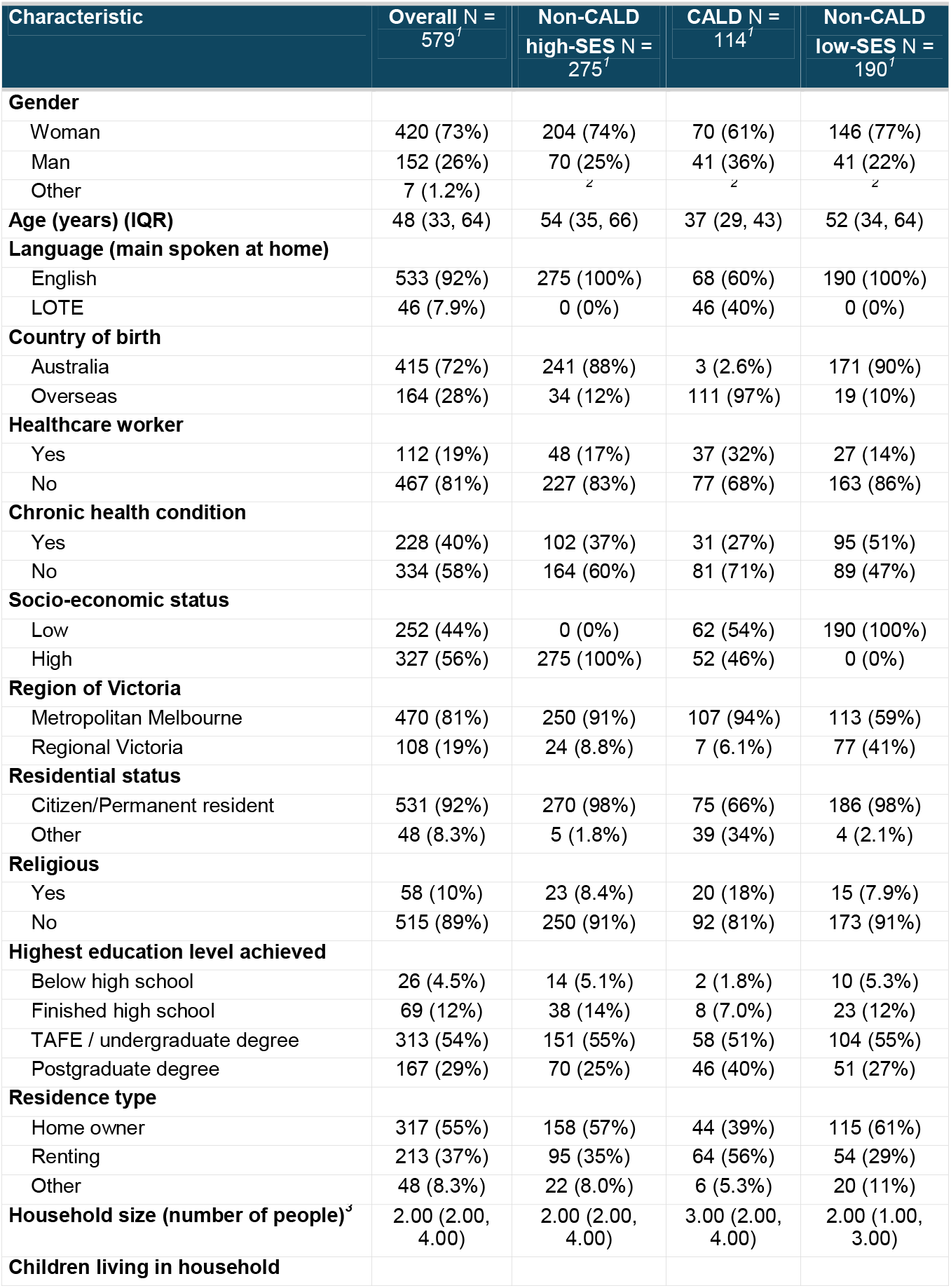
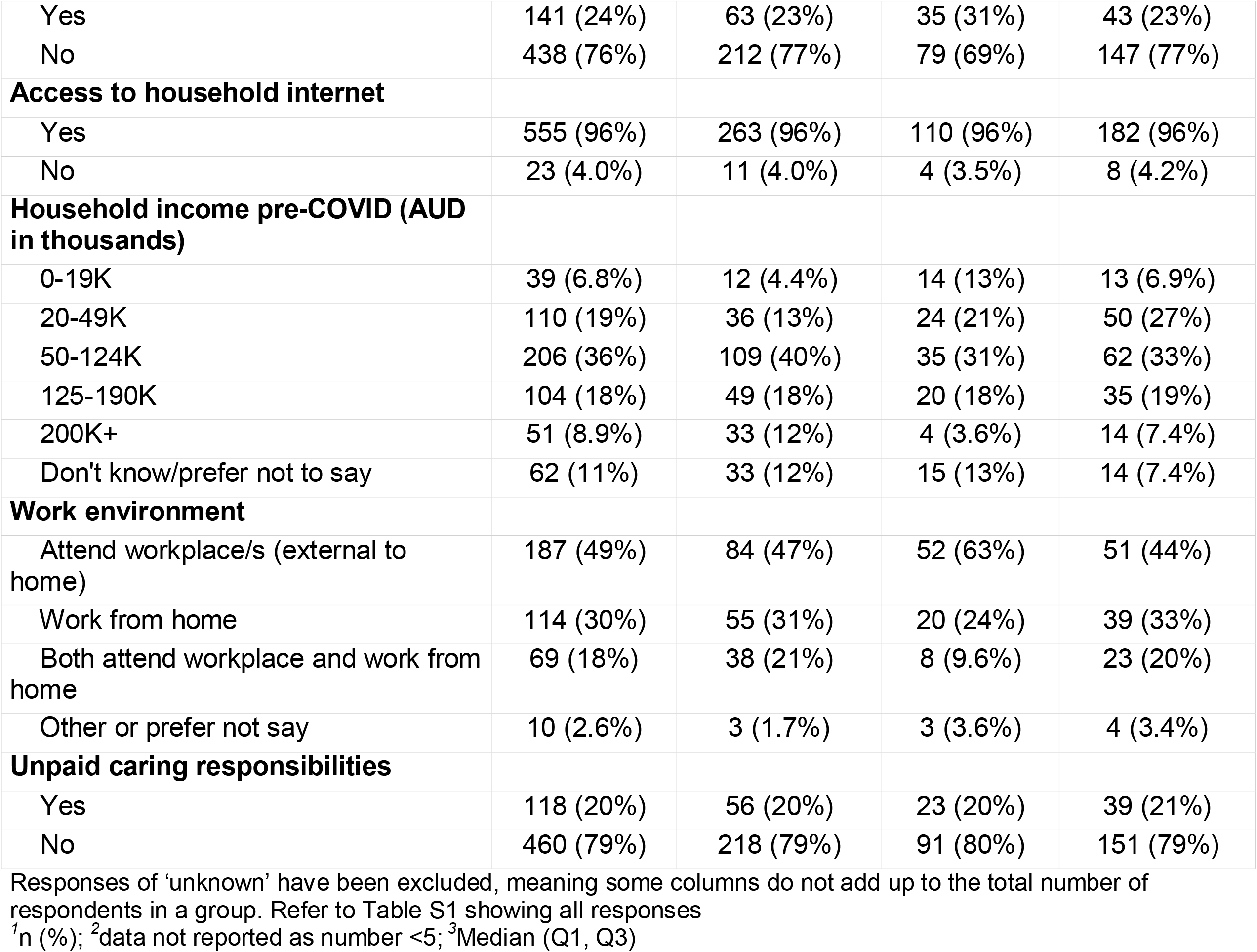
Sociodemographic characteristics of Optimise study participants at baseline survey, by exposure group (N=579)

The CALD group were younger (median age 37 years), and a higher proportion were healthcare workers (n=37, 32%) compared to ‘non-CALD high-SES’ and ‘non-CALD low-SES’ groups. A greater proportion of ‘non-CALD low-SES’ had a chronic health condition (n=95, 51%) and lived in regional Victoria (n=77, 41%) compared to the other two groups. The CALD group had a larger household size (median of 3 people), were less likely to be permanent residents of Australia (n=75, 66%), and a higher proportion had children living in the household (n=35, 31%) compared to the other two groups. A higher proportion of the CALD group attended workplaces outside of the home (n=52, 63%) compared to the other two groups.

### Adherence to public health Recommendations and Restrictions

Across all three groups, participants adopted more risk reduction behaviours during the lockdown period compared to the non-lockdown period (**Table 3**). Within the lockdown period, there was a statistically significant difference between groups except for two measures, ‘disinfecting surfaces and objects’ and ‘avoiding seeing people who are older/vulnerable’. Nevertheless, adherence to avoiding seeing older/vulnerable people was still high (>70% in each group) with the CALD group having the highest adherence (n=70, 90%).

**Table 3.**
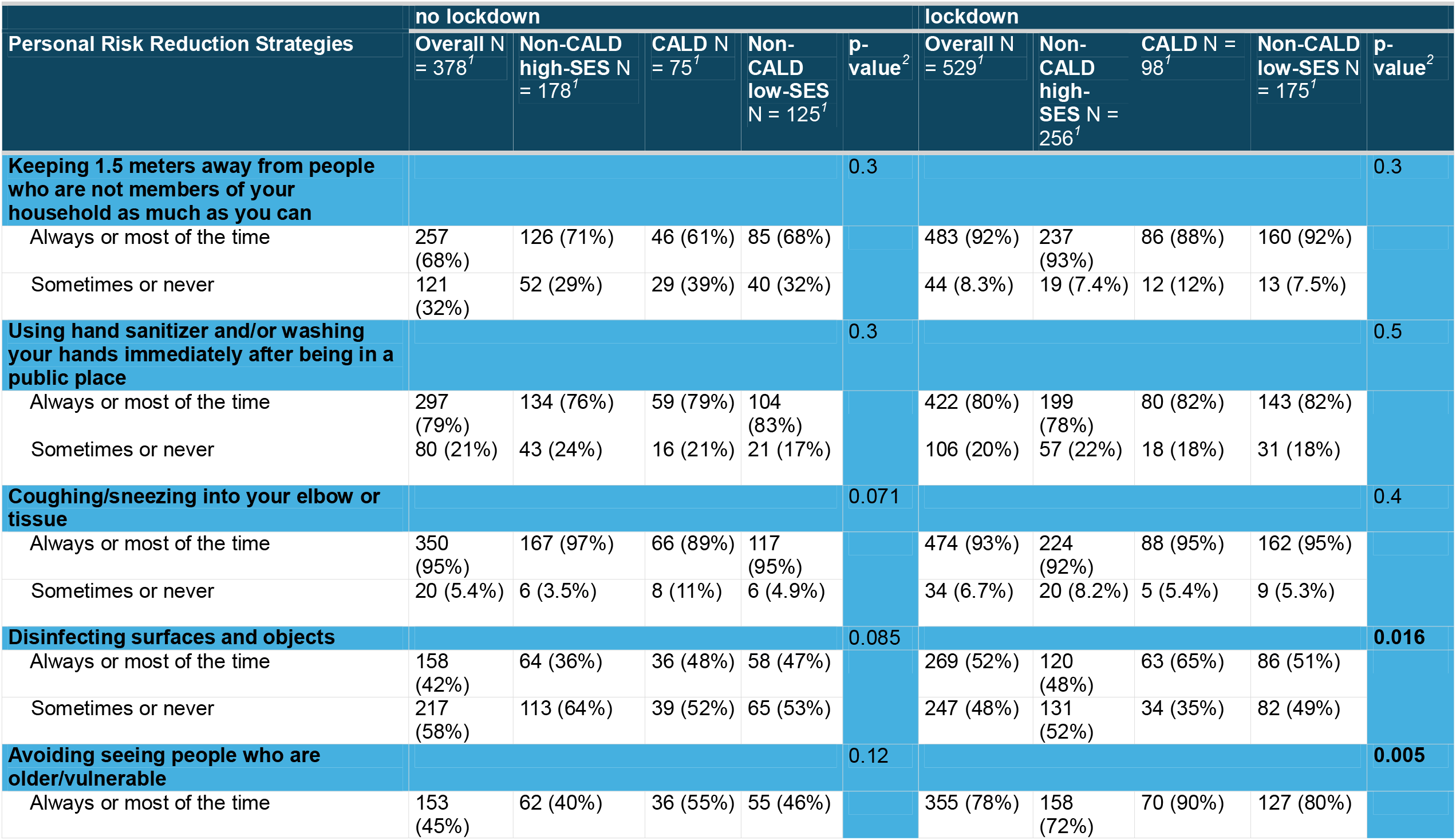
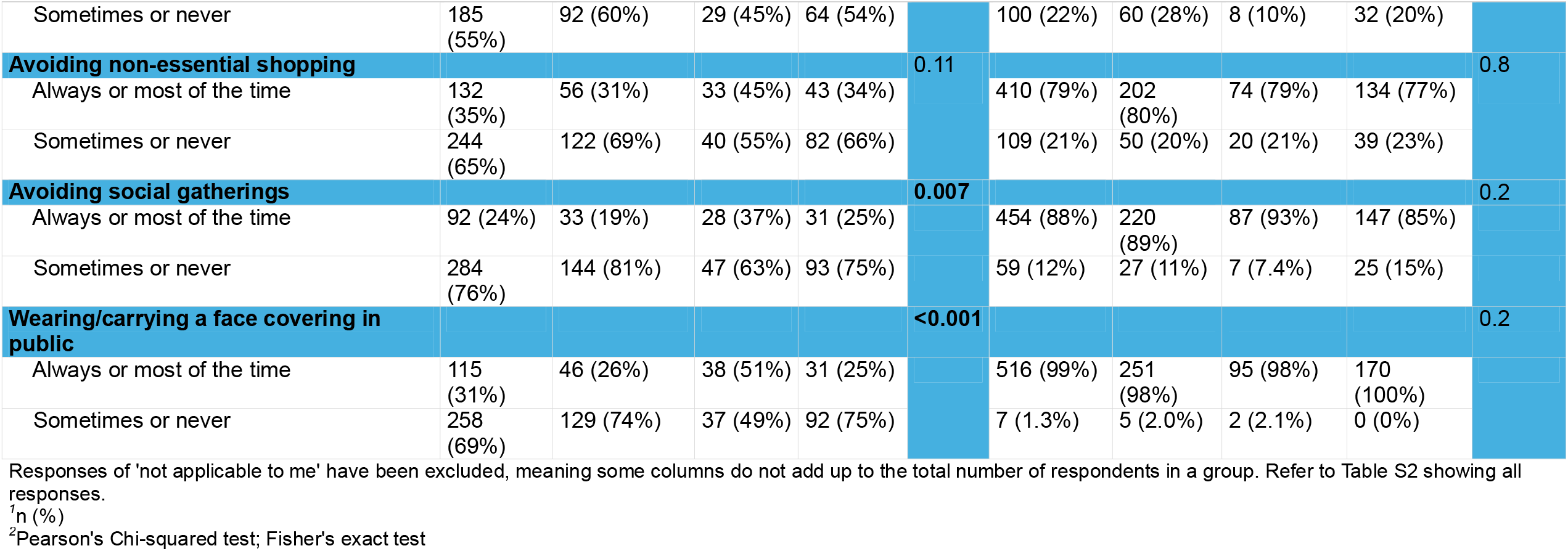
Adherence to Restrictions and Recommendations by group during lockdown and non-lockdown periods.

During the non-lockdown period, there was no statistical difference between groups except for two measures, ‘avoiding social gatherings’ and ‘wearing/carrying a face covering’. The CALD group had the highest adherence to these public health measures (avoiding gatherings [n=28, 37%] and wearing/carrying a face covering [n=46, 26%] compared to ‘non-CALD high-SES’ and ‘non-CALD low-SES’ groups.

### Adherence to public health Requirements

Of the 132 individuals who reported testing for COVID-19 during either study periods, 37 (28%) reported leaving home while awaiting the COVID-19 test result (**Table 4**). Among participants who reported testing for COVID, 44% (14/32) of CALD participants reported leaving home, compared to 14% (6/14) of ‘non-CALD low-SES’ and 29% (17/58) of the ‘non-CALD high-SES’ group. Of the participants who left home while awaiting a test, 68% (23/32) worked outside of the home and 18% (6/32) were working from home or not working. In the group who remained at home while awaiting a test result, 40% (36/95) worked outside of the home, and 42% (38/95) worked from home or were not working (**Table 4**). The most common reasons for leaving home included for work, for essential food or medications, for exercise and for medical appointments (**Table S3**).

**Table 4.** Adherence to Requirements and characteristics for N=132 participants who reported testing for COVID-19 in the past 7 days.

| Characteristic | Left home before receiving a result N = 37<sup>1</sup> | Did not leave home before receiving a result N = 95<sup>1</sup> |
| --- | --- | --- |
| **Population group** |  |  |
| Non-CALD high-SES | 17 (29%) | 41 (71%) |
| CALD | 14 (44%) | 18 (56%) |
| Non-CALD low-SES | 6 (14%) | 36 (86%) |
| **Baseline characteristics** |  |  |
| **Gender** |  |  |
| Man | 15 (41%) | 21 (22%) |
| Woman | 22 (59%) | 73 (77%) |
| Non-binary | 0 (0%) | <sup>2</sup> |
| **Children living in the household** |  |  |
| Yes | 10 (27%) | 21 (22%) |
| No | 27 (73%) | 74 (78%) |
| **Information from follow-up survey completed close to COVID-19 test** |  |  |
| **Work environment in past 4 weeks** |  |  |
| Attend workplace/s (external to home) | 23 (68%) | 36 (40%) |
| Both attend workplace and work from home | 5 (15%) | 16 (18%) |
| Work from home | 0 (0%) | 10 (11%) |
| Not working | 6 (18%) | 28 (31%) |
| Missing (Did not complete a follow-up survey) | 3 | 5 |
| **Unpaid caring responsibilities in past 4 weeks** |  |  |
| Yes | 7 (21%) | 23 (26%) |
| No | 27 (79%) | 66 (73%) |
| Prefer not to say | 0 (0%) | 1 (1.1%) |
| Missing (Did not complete a follow-up survey) | 3 | 5 |

<sup>1</sup>n (%) <sup>2</sup>data not reported as number <5

### Sub-group analysis

There was a trend toward CALD-low-SES behaving similarly to the non-CALD low-SES group, and CALD-high-SES behaving similarly to the non-CALD high-SES group (**Table S2**). However, the numbers in each of these groups were small (n<51) and the differences between groups was not statistically significant (p≥0.05).

## DISCUSSION

Data from our Victorian study shows high adherence (>85%) to public health measures during the lockdown period across all three exposure groups, suggesting widespread acceptance of the need for behavioural strategies to reduce COVID-19 transmission. Some behaviours, like hand sanitiser use and coughing into the elbow remained high during non-lockdown periods, suggesting a more sustained change compared to measures like mask use. However, recommendations and restrictions varied in their impact on personal freedoms and social connections. Our study shows that recommendations that were easy to implement individually and had minimal social disruption were more consistently followed, while those limiting social interaction – such as avoiding older/vulnerable people, non-essential shopping, and social gatherings – were discontinued during non-lockdown periods. Recognising the personal impacts different risk reduction strategies have on individuals is essential for understanding non-adherence, as future governments may have limited social license for more restrictive measures.

The CALD and non-CALD, low-SES groups in this study had higher uptake of some recommendations and restrictions compared with the high-SES, non-CALD groups, during lockdown and non-lockdown periods. This trend was also observed in the sub-analysis where there was greater adoption of public health measures among the low-SES group, regardless of CALD status, compared to the high-SES group. This finding challenges the view that the higher incidence of COVID-19 infection and mortality in CALD groups and those living in low-SES areas was attributable to lower uptake of behavioural health measures.^15^

One quarter of participants awaiting COVID-19 test results – a period of high transmission risk – did not stay at home, with the CALD group representing the highest proportion. This finding contrasts with their otherwise high adherence to recommendations and restrictions, suggesting that while CALD participants understood the importance of behavioural measures, compliance was not always feasible. Compared to the other groups, the CALD group included more younger people, healthcare workers, larger households, working outside of the home, and those with children. Fewer held citizenship/permanent residency, potentially limiting Medicare access.

‘Work’ was the commonest reason for participants to leave home while awaiting a COVID-19 test, implying that despite government support of COVID-19 leave, participants could not take time off work. Recent migrants and foreign students often occupy lower paid casual jobs in the service industry or gig economy, increasing their exposure to SARS-CoV-2 and economic vulnerabilities.^16^ Factors such as larger households, extended family living, essential worker status, lower health literacy, and limited healthcare access, have been shown to contribute to higher COVID-19 infection rates.^17^ These factors may explain the higher rate of non-adherence during COVID-19 isolation for the CALD group.

Our findings highlight the challenges individuals faced in adhering to public health requirements that limited their ability to perform essential daily tasks. This contrasts with high adherence to other behavioural changes that did not have the same social and economic cost. These findings emphasise the need for more nuanced and tailored support when implementing public health measures, particularly those that require individuals to sacrifice social or financial responsibilities in order to comply. A 2025 Australian Human Rights Commission report highlighted the individual nature of pandemic experiences in Australia, emphasising that ‘one size does not fit all’ when supporting those affected by emergency responses, as needs vary greatly depending on personal and household circumstances.^18^

### Limitations

This study has several limitations. The IRSD is a well-described proxy for socio-economic status in Australia,^19-21^ however postcode-level data may not reflect the socio-economic circumstances of individual households. While some participants’ socio-economic profile may differ from their IRSD classification, the large sample size of our cohort likely minimised the impact on our findings.

Secondly, our data may be limited by recall and social desirability bias. The recall bias is likely mitigated by the contemporaneous nature of the observations, with a recall period of seven days for the diaries and a recall period of 28 days for the surveys. We reduced social desirability bias by anonymising data reporting, providing self-administered questionnaires and avoiding leading questions.

Despite these limitations the study leverages a large, longitudinal observational cohort that is unique in the Australian context, with high retention and contemporaneous data collection sustained throughout the duration of the pandemic emergency health measures and beyond.^22^

## Conclusion

Our study highlights the overall high adherence to public health measures during the lockdown period examined in Victoria. CALD individuals had a higher rate of adherence to public recommendations and restrictions than the non-CALD, high-SES population, although they reported higher rates of lower adoption to requirements while in isolation, a time of significant COVID-19 transmission risk. Factors such as occupation as an essential worker, larger household sizes, living with children, and decreased access to Australia’s public healthcare system may contribute to the non-adherence. Our findings suggest that adherence to public health measures that require individuals to sacrifice social or financial responsibilities in order to comply, require tailored support during future pandemics to address the diverse circumstances of priority populations.

## Data Availability

All data produced in the present study are available upon reasonable request to the authors

## Acknowledgements

Optimise is a partnership between Burnet Institute and Peter Doherty Institute in collaboration with University of Melbourne, Swinburne University of Technology, Monash University, La Trobe University, Murdoch Children’s Research Institute, the Centre for Culture Ethnicity and Health, and the Health Issues Centre. The authors gratefully acknowledge the generosity of the community members who participated in the study. The authors appreciatively acknowledge the work of all Optimise project team members and collaborators who have contributed to the ongoing delivery of the study. The authors gratefully acknowledge the contribution to this work of the Victorian Operational Infrastructure Support Program received by the Burnet Institute.

