## Supplemental Tables for "Adherence to Public Health Recommendations, Restrictions, and Requirements among Priority Populations at Risk for COVID-19 Mortality and Infection in Australia"

**SUPPLEMENTARY MATERIAL**

| **Table S1**: Sociodemographic characteristics of participants, unknown responses included | Page 2 |
| --- | --- |
| **Table S2**: Adherence to Restrictions and Recommendations by group during lockdown and non-lockdown periods, with CALD divided into low- and high- SES | Page 5 |
| **Table S3:** Reasons provided for leaving home while awaiting the results of a COVID-19 test | Page 6 |

**Table S1: Sociodemographic characteristics of participants, unknown responses included**

| **Characteristic** | **Overall** N = 579*^1^* | **Non-CALD high-SES** N = 275*^1^* | **CALD** N = 114*^1^* | **Non-CALD low-SES** N = 190*^1^* |
| --- | --- | --- | --- | --- |
| **Gender** |  |  |  |  |
| Woman | 420 (73%) | 204 (74%) | 70 (61%) | 146 (77%) |
| Man | 152 (26%) | 70 (25%) | 41 (36%) | 41 (22%) |
| Other | 7 (1.2%) | 1 (0.4%) | 3 (2.6%) | 3 (1.6%) |
| **Age** | 48 (33, 64) | 54 (35, 66) | 37 (29, 43) | 52 (34, 64) |
| **Language** |  |  |  |  |
| English | 533 (92%) | 275 (100%) | 68 (60%) | 190 (100%) |
| LOTE | 46 (7.9%) | 0 (0%) | 46 (40%) | 0 (0%) |
| **Country of birth** |  |  |  |  |
| Australia | 415 (72%) | 241 (88%) | 3 (2.6%) | 171 (90%) |
| Overseas | 164 (28%) | 34 (12%) | 111 (97%) | 19 (10%) |
| **Healthcare worker** |  |  |  |  |
| Yes | 112 (19%) | 48 (17%) | 37 (32%) | 27 (14%) |
| No | 467 (81%) | 227 (83%) | 77 (68%) | 163 (86%) |
| **Chronic health condition** |  |  |  |  |
| Yes | 228 (40%) | 102 (37%) | 31 (27%) | 95 (51%) |
| No | 334 (58%) | 164 (60%) | 81 (71%) | 89 (47%) |
| Other | 15 (2.6%) | 9 (3.3%) | 2 (1.8%) | 4 (2.1%) |
| Unknown | 2 (0.3%) | 0 (0%) | 0 (0%) | 2 (1%) |
| **Socio-economic status** |  |  |  |  |
| Low | 252 (44%) | 0 (0%) | 62 (54%) | 190 (100%) |
| High | 327 (56%) | 275 (100%) | 52 (46%) | 0 (0%) |
| **Region of Victoria** |  |  |  |  |
| Metropolitan Melbourne | 470 (81%) | 250 (91%) | 107 (94%) | 113 (59%) |
| Regional Victoria | 108 (19%) | 24 (8.8%) | 7 (6.1%) | 77 (41%) |
| Unknown | 1 (0.2%) | 1 (0.4%) | 0 (0%) | 0 (0%) |
| **Residential status** |  |  |  |  |
| Permanent resident | 531 (92%) | 270 (98%) | 75 (66%) | 186 (98%) |
| Other | 48 (8.3%) | 5 (1.8%) | 39 (34%) | 4 (2.1%) |
| **Religious** |  |  |  |  |
| Yes | 58 (10%) | 23 (8.4%) | 20 (18%) | 15 (7.9%) |
| No | 515 (89%) | 250 (91%) | 92 (81%) | 173 (91%) |
| Prefer not to say | 6 (1.0%) | 2 (0.7%) | 2 (1.8%) | 2 (1.1%) |
| **Highest education level achieved** |  |  |  |  |
| Below highschool | 26 (4.5%) | 14 (5.1%) | 2 (1.8%) | 10 (5.3%) |
| Finished highschool | 69 (12%) | 38 (14%) | 8 (7.0%) | 23 (12%) |
| TAFE / undergraduate degree | 313 (54%) | 151 (55%) | 58 (51%) | 104 (55%) |
| Postgraduate degree | 167 (29%) | 70 (25%) | 46 (40%) | 51 (27%) |
| Prefer not to say | 4 (0.7%) | 2 (0.7%) | 0 (0%) | 2 (1.1%) |
| **Residence type** |  |  |  |  |
| Home owner | 317 (55%) | 158 (57%) | 44 (39%) | 115 (61%) |
| Renting | 213 (37%) | 95 (35%) | 64 (56%) | 54 (29%) |
| Other | 48 (8.3%) | 22 (8.0%) | 6 (5.3%) | 20 (11%) |
| Unknown | 1 (0.2%) | 0 (0%) | 0 (0%) | 1 (0.5%) |
| **Household size** | 2.00 (2.00, 4.00) | 2.00 (2.00, 4.00) | 3.00 (2.00, 4.00) | 2.00 (1.00, 3.00) |
| **Children living in household** |  |  |  |  |
| Yes | 141 (24%) | 63 (23%) | 35 (31%) | 43 (23%) |
| No | 438 (76%) | 212 (77%) | 79 (69%) | 147 (77%) |
| **Access to household internet** |  |  |  |  |
| Yes | 555 (96%) | 263 (96%) | 110 (96%) | 182 (96%) |
| No | 23 (4.0%) | 11 (4.0%) | 4 (3.5%) | 8 (4.2%) |
| Don't know | 1 (0.2%) | 1 (0.4%) | 0 (0%) | 0 (0%) |
| **Household income pre-COVID** |  |  |  |  |
| 0-19K | 39 (6.8%) | 12 (4.4%) | 14 (13%) | 13 (6.9%) |
| 20-49K | 110 (19%) | 36 (13%) | 24 (21%) | 50 (27%) |
| 50-124K | 206 (36%) | 109 (40%) | 35 (31%) | 62 (33%) |
| 125-190K | 104 (18%) | 49 (18%) | 20 (18%) | 35 (19%) |
| 200k+ | 51 (8.9%) | 33 (12%) | 4 (3.6%) | 14 (7.4%) |
| Don't know/prefer not to say | 62 (11%) | 33 (12%) | 15 (13%) | 14 (7.4%) |
| Unknown | 7 (1.2%) | 3 (1.1%) | 2 (1.8%) | 2 (1%) |
| **Work environment at baseline** |  |  |  |  |
| Attend a single workplace | 127 (33%) | 57 (32%) | 33 (40%) | 37 (32%) |
| Attend multiple workplaces | 60 (16%) | 27 (15%) | 19 (23%) | 14 (12%) |
| Both workplace and work from home | 69 (18%) | 38 (21%) | 8 (9.6%) | 23 (20%) |
| Work from home | 114 (30%) | 55 (31%) | 20 (24%) | 39 (33%) |
| Other or prefer not say | 10 (2.6%) | 3 (1.7%) | 3 (3.6%) | 4 (3.4%) |
| Not working | 199 (34%) | 95 (24%) | 31 (27%) | 73 (38%) |
| **Unpaid caring responsibilities at baseline** |  |  |  |  |
| Yes | 118 (20%) | 56 (20%) | 23 (20%) | 39 (21%) |
| No | 460 (79%) | 218 (79%) | 91 (80%) | 151 (79%) |
| Don't know | 1 (0.2%) | 1 (0.4%) | 0 (0%) | 0 (0%) |
| *^1^*n (%); Median (Q1, Q3) | | | | |

**Table S2: Adherence to Restrictions and Recommendations by group during lockdown and non-lockdown periods, with CALD divided into low- and high- SES**

|  | **no lockdown** | | | | | | **lockdown** | | | | | |
| --- | --- | --- | --- | --- | --- | --- | --- | --- | --- | --- | --- | --- |
| **Characteristic** | **Overall** N = 378*^1^* | **Non-CALD high-SES**  N = 178*^1^* | **Non-CALD low- SES** N = 125*^1^* | **CALD- high- SES** N = 40*^1^* | **CALD-low SES** N = 35*^1^* | **p-value***^2^* | **Overall** N = 529*^1^* | **Non-CALD high-SES**  N = 256*^1^* | **Non-CALD low- SES** N = 175*^1^* | **CALD-high- SES** N = 47*^1^* | **CALD-low- SES** N = 51*^1^* | **p-value***^3^* |
| **Keeping 1.5 metres away from people who are not members of your household as much as you can** |  |  |  |  |  | 0.5 |  |  |  |  |  | 0.2 |
| Always or most of the time | 257 (68%) | 126 (71%) | 85 (68%) | 24 (60%) | 22 (63%) |  | 483 (91%) | 237 (93%) | 160 (91%) | 39 (83%) | 47 (92%) |  |
| Sometimes or never | 121 (32%) | 52 (29%) | 40 (32%) | 16 (40%) | 13 (37%) |  | 44 (8.3%) | 19 (7.4%) | 13 (7.4%) | 8 (17%) | 4 (7.8%) |  |
| Not applicable to me | 0 (0%) | 0 (0%) | 0 (0%) | 0 (0%) | 0 (0%) |  | 2 (0.4%) | 0 (0%) | 2 (1.1%) | 0 (0%) | 0 (0%) |  |
| **Wearing or carrying a face covering in public** |  |  |  |  |  | **0.001** |  |  |  |  |  | **0.018** |
| Always or most of the time | 115 (30%) | 46 (26%) | 31 (25%) | 17 (43%) | 21 (60%) |  | 516 (98%) | 251 (98%) | 170 (97%) | 45 (96%) | 50 (98%) |  |
| Sometimes or never | 258 (68%) | 129 (72%) | 92 (74%) | 23 (58%) | 14 (40%) |  | 7 (1.3%) | 5 (2.0%) | 0 (0%) | 1 (2.1%) | 1 (2.0%) |  |
| Not applicable to me | 5 (1.3%) | 3 (1.7%) | 2 (1.6%) | 0 (0%) | 0 (0%) |  | 6 (1.1%) | 0 (0%) | 5 (2.9%) | 1 (2.1%) | 0 (0%) |  |
| **Using hand sanitizer and/or washing your hands immediately after being in a public place** |  |  |  |  |  | 0.2 |  |  |  |  |  | 0.6 |
| Always or most of the time | 297 (79%) | 134 (75%) | 104 (83%) | 35 (88%) | 24 (69%) |  | 422 (80%) | 199 (78%) | 143 (82%) | 37 (79%) | 43 (84%) |  |
| Sometimes or never | 80 (21%) | 43 (24%) | 21 (17%) | 5 (13%) | 11 (31%) |  | 106 (20%) | 57 (22%) | 31 (18%) | 10 (21%) | 8 (16%) |  |
| Not applicable to me | 1 (0.3%) | 1 (0.6%) | 0 (0%) | 0 (0%) | 0 (0%) |  | 1 (0.2%) | 0 (0%) | 1 (0.6%) | 0 (0%) | 0 (0%) |  |
| **Coughing/sneezing into your elbow or tissue** |  |  |  |  |  | 0.2 |  |  |  |  |  | 0.2 |
| Always or most of the time | 350 (93%) | 167 (94%) | 117 (94%) | 33 (83%) | 33 (94%) |  | 474 (90%) | 224 (88%) | 162 (93%) | 39 (83%) | 49 (96%) |  |
| Sometimes or never | 20 (5.3%) | 6 (3.4%) | 6 (4.8%) | 6 (15%) | 2 (5.7%) |  | 34 (6.4%) | 20 (7.8%) | 9 (5.1%) | 4 (8.5%) | 1 (2.0%) |  |
| Not applicable to me | 8 (2.1%) | 5 (2.8%) | 2 (1.6%) | 1 (2.5%) | 0 (0%) |  | 21 (4.0%) | 12 (4.7%) | 4 (2.3%) | 4 (8.5%) | 1 (2.0%) |  |
| **Disinfecting surfaces and objects** |  |  |  |  |  | 0.3 |  |  |  |  |  | 0.068 |
| Always or most of the time | 158 (42%) | 64 (36%) | 58 (46%) | 18 (45%) | 18 (51%) |  | 269 (51%) | 120 (47%) | 86 (49%) | 29 (62%) | 34 (67%) |  |
| Sometimes or never | 217 (57%) | 113 (63%) | 65 (52%) | 22 (55%) | 17 (49%) |  | 247 (47%) | 131 (51%) | 82 (47%) | 17 (36%) | 17 (33%) |  |
| Not applicable to me | 3 (0.8%) | 1 (0.6%) | 2 (1.6%) | 0 (0%) | 0 (0%) |  | 13 (2.5%) | 5 (2.0%) | 7 (4.0%) | 1 (2.1%) | 0 (0%) |  |
| **Avoiding seeing people who are older/vulnerable** |  |  |  |  |  | 0.12 |  |  |  |  |  | **0.006** |
| Always or most of the time | 153 (40%) | 62 (35%) | 55 (44%) | 19 (48%) | 17 (49%) |  | 355 (67%) | 158 (62%) | 127 (73%) | 32 (68%) | 38 (75%) |  |
| Sometimes or never | 185 (49%) | 92 (52%) | 64 (51%) | 18 (45%) | 11 (31%) |  | 100 (19%) | 60 (23%) | 32 (18%) | 6 (13%) | 2 (3.9%) |  |
| Not applicable to me | 40 (11%) | 24 (13%) | 6 (4.8%) | 3 (7.5%) | 7 (20%) |  | 74 (14%) | 38 (15%) | 16 (9.1%) | 9 (19%) | 11 (22%) |  |
| **Avoiding non-essential shopping** |  |  |  |  |  | **0.050** |  |  |  |  |  | 0.5 |
| Always or most of the time | 132 (35%) | 56 (31%) | 43 (34%) | 18 (45%) | 15 (43%) |  | 410 (78%) | 202 (79%) | 134 (77%) | 34 (72%) | 40 (78%) |  |
| Sometimes or never | 244 (65%) | 122 (69%) | 82 (66%) | 21 (53%) | 19 (54%) |  | 109 (21%) | 50 (20%) | 39 (22%) | 11 (23%) | 9 (18%) |  |
| Not applicable to me | 2 (0.5%) | 0 (0%) | 0 (0%) | 1 (2.5%) | 1 (2.9%) |  | 10 (1.9%) | 4 (1.6%) | 2 (1.1%) | 2 (4.3%) | 2 (3.9%) |  |
| **Avoiding social gatherings** |  |  |  |  |  | **0.049** |  |  |  |  |  | 0.091 |
| Always or most of the time | 92 (24%) | 33 (19%) | 31 (25%) | 15 (38%) | 13 (37%) |  | 454 (86%) | 220 (86%) | 147 (84%) | 38 (81%) | 49 (96%) |  |
| Sometimes or never | 284 (75%) | 144 (81%) | 93 (74%) | 25 (63%) | 22 (63%) |  | 59 (11%) | 27 (11%) | 25 (14%) | 6 (13%) | 1 (2.0%) |  |
| Not applicable to me | 2 (0.5%) | 1 (0.6%) | 1 (0.8%) | 0 (0%) | 0 (0%) |  | 16 (3.0%) | 9 (3.5%) | 3 (1.7%) | 3 (6.4%) | 1 (2.0%) |  |
| *^1^*n (%) | | | | | | | | | | | | |
| *^2^*Fisher's exact test | | | | | | | | | | | | |
| *^3^*Fisher's exact test; Pearson's Chi-squared test | | | | | | | | | | | | |

**Table S3: Reasons provided for leaving home while awaiting the results of a COVID-19 test**

| **Reason** | **Lockdown** | **No lockdown** | **Total** |
| --- | --- | --- | --- |
| Work | 19 | 7 | 26 |
| Essential food | 7 | 4 | 11 |
| Exercise | 6 | 3 | 9 |
| Medical care | 3 | 0 | 3 |
| Essential medications | 0 | 2 | 2 |
| Pick up/drop off a household member from work, school, childcare, appointments | 2 | 2 | 4 |
| Emergency situation | 1 | 0 | 1 |
| Visit family, friends or partner | 1 | 1 | 2 |
| Other | 5 | 1 | 6 |
